# Modeling the COVID-19 transmission in Italy: The roles of asymptomatic cases, social distancing, and lockdowns in the first and the second waves

**DOI:** 10.1101/2021.01.08.21249273

**Authors:** Muhamad Khairulbahri

## Abstract

The SEIR model of COVID-19 is developed to investigate the roles of physical distancing, lockdowns and asymptomatic cases in Italy. In doing so, two types of policies including behavioral measures and lockdown measures are embedded in the model. Compared with existing models, the model successfully reproduces similar multiple observed outputs such as infected and recovered patients in Italy by July 2020. This study concludes that the first policy is important once the number of infected cases is relatively low. However, once the number of infected cases is very high so the society cannot identify infected and disinfected people, the second policy must be applied soon. It is thus this study suggests that relaxed lockdowns lead to the second wave of the COVID-19 around the world. It is hoped that the model can enhance our understanding on the roles of behavioral measures, lockdowns, and undocumented cases, so-called asymptomatic cases, on the COVID-19 flow.

## I. Introduction

Despite its high quality of the national healthcare program, Italy was the epicenter of the corona virus disease (the COVID-19) during the European first wave. The Italy first closure owing to COVID-19 was on February 22nd, 2020. As COVID-case spiked, the government started the first measure on March 4th by closing public places such as stores, and schools as well as enforcing physical distancing in severely affected areas close to Milan and Venice (Sjödin et al., 2020). This lockdown was lengthened on March 9th, owing to a rise of documented cases and confirmed deaths at about 7,000 people and 400 people respectively.

Some existing studies discussed issues relate to Italian COVID-19 cases such as policy measures (Flaxman et al., 2020; Lavezzo et al., 2020; Sjödin et al., 2020), the dynamics of high COVID-19 cases (De Natale et al., 2020; Garazzino et al., 2020; Gatto et al., 2020; Goumenou et al., 2020; Grasselli et al., 2020; Grossi et al., 2020), and comparisons of the COVID-19 in Italy and China (Nesteruk, 2020). It appears that existing studies successfully has modelled the COVID-19 cases in Italy and analyzed the main factors of Italian COVID-19 flow. However, at the best author knowledge, no available study has successfully modelled two different notified policies in tackling the COVID-19: *the behavior policy and the lockdown policy*. This is important as other studies (Carlucci et al., 2020; Graffigna et l., 2020; Meier et al., 2020; Pagnini et al., 2020) highlighted the importance the first and the second policies in minimizing the COVID-19 flow in Italy. Hence, this study compares the efficacy two different measures in the Italian COVID-19 cases.

This study also investigates three important cases simultaneously including recovered, infected, and dead patients, especially during the first wave in Italy. As some studies mentioned (De Natale et al., 2020; Flaxman et al., 2020; Gatto et al., 2020; Lavezzo et al., 2020) possible roles of undocumented cases, this study also investigates the role of the undocumented cases in Italy. With keep these points in mind, the author believes that modeling the COVID-19 in Italy can give us understanding the dynamics flow of the COVID-19 in terms of behavior, lockdown policies, and undocumented cases.

Moreover, this study offers more comprehensive analysis as this study analyses the quantitative impacts of undocumented cases, the quantitative impacts of behavioral measures, and the quantitative impacts of lockdowns than existing studies (e.g., Dehning et al., 2020; Raheem, 2020; Sjödin et al., 2020).

In the first section, this study explains existing studies of COVID-19 in Italy. Furthermore, this study explains the system dynamics approach to understand dynamic of susceptible, exposed, infectious and recovered patients in Italy. The system dynamics approach has a long tradition to model health issues such as epidemiology, healthcare facilities, and non-communicable diseases (Darabi, N. & Hosseinichimeh, 2020; Homer, & Hirsch, 2006). Because of this, this study develops a SEIR model based on the system dynamics approach to simulate deaths, recoveries, and incidences in Italy.

## II. Data and methods

Required data were collected from https://www.worldometers.info/coronavirus/, www.rki.de (Robert Koch Institute) and https://coronavirus.jhu.edu (John Hopkins University). Three data types including active cases (infected people), deaths and recovered people were extracted from aforementioned sources.

Previous studies based on the system dynamics approach have successfully simulated infectious, non-infectious diseases, and other healthcare issues (Donsimoni et al., 2020; Darabi & Hosseinichimeh, 2020; Davahli et al., 2020; Homer & Hirsch, 2006). Owing to this, the SEIR model used in this study is developed based on the system dynamics approach. Despite usefulness of the SEIR model, some uncertain parameters have not been defined or unknown.

To obtain the best parameter values, this study applies the Markov Chain Monte Carlo (MCMC) calibration process owing to unknown parameters using built functions in Vensim©.

The model starts the simulation from January 1^st^, 2020 and runs in daily terms until the end of July 2020. Simulated parameters such as an incubation time, Ro, and an infection duration are based on existing studies. It is important to note that the SEIR model will be set in two types of the first confirmed cases. Previous studies (e.g., Usuelli, 2020) noted that COVID-19 models of Italy should assume that the first confirmed case is infected Chinese tourists who was acknowledged as an imported COVID-19 case on January 31^st^, 2020. While other studies (De Natale et al., 2020; Gatto et al., 2020; 2020; Sjödin et al., 2020) assumed that the first Italian confirmed case should be the first infected male on February 21^st^, 2020.

By comparing two types of the first confirmed cases, this study wants to prove the importance of behavioral measures such as physical distancing and handwashing. The logic is if there is no much differences of estimated parameter values between the two first confirmed cases, this intuitively means that behavioral measures was effective in hindering the COVID-19 flow from January 31^st^, 2020 to February 21^st^, 2020.

## III. Discussion and results

To obtain the best value for each parameter, at the first step, the SDM is embedded by range values stated in table 1 and at second step, we run the Markov Chain Monte Carlo (MCMC) calibration available in Vensim©. To accommodate uncertain parameter values, this study set range values based on existing studies. For instance, as other studies (Lauer et al., 2020; Yu et al., 2020), this study sets Ro ranges between 2.4 and 3.7.

**Table 1.** Parameter values of the SDM. Bracketed values are min and max values based on existing studies

| No | Names | Values | References |
| --- | --- | --- | --- |
| 1 | The **two** first confirmed cases | January 31<sup>st</sup>, 2020<br>February 21<sup>st</sup>, 2020 | (De Natale et al., 2020; Goumenou et al., 2020; Raheem, 2020) |
| 2 | Ro (basic reproduction number) | 3.5 (3-4) | (Garazzzino et al., 2020; ISSc, 2020) |
| 3 | Incubation time | 4 days (3-5) days | (Lauer et al., 2020; Sjödin et al., 2020; Yu et al., 2020) |
| 4 | Infection duration | 26 (7-45) days | (ISSc, 2020) |
| 5 | Recovery time | 8 (5-12) days | (Grasselli et al., 2020) |
| 6 | Behavioral reaction time<br>The behavioral reduction time is measured between the first infection case and the beginning of the behavioral risk reduction actions. This time measurement also applies to other time measurements. | 15 days on March 4<sup>th</sup> (public place closures) and 22<sup>nd</sup> (the national lockdown)<br>There was also the lockdown on March 8<sup>th</sup> (The lockdown of Northern Italy) | (Goumenou et al., 2020; ISSc, 2020) |
| 7 | Behavioral risk reduction | This policy is voluntary acts without legal enforcement.<br>This is assumed to be between 10% and 50% |  |
| 8 | Lockdown reaction time<br>As there are many lockdowns, another variable i.e., “delay time” (1-5 days) is inserted in the SEIR model. | 45 days (after the first case) | (Garazzzino et al., 2020; ISSc, 2020) |
| 9 | lockdown risk reduction | The lockdowns are very strict and enforced by law, so the efficacy is high (60%-95%) |  |
| 10 | Fraction of undocumented cases upon documented cases | 55% (46-62) % | Li et al. (2020)<br>Flaxman et al. (2020); Gatto et al. (2020); Lavezzo et al. (2020) |

### The SEIR model

The system dynamics model (SDM), so called stock-flow model, for Italian cases can be seen in figure 1. The SDM consists of five stocks such as the susceptible, infected, and recovered cases. The SDM also accommodates the impacts of behavioral measures such as physical distancing, hand washing, and mask covers. For these measures, the SDM applies two variables namely, “*behavioral risk reduction*” and “*behavioral reaction time*”. These two variables respectively explain efficacy of and starting time of aforementioned measures in minimizing the flow of COVID-19. The more striking COVID-19 cases, the more critical or defensive measures are taken such as isolating patients and lockdowns. In considering striking cases, the SDM also captures the impacts of lockdowns through two variables: “*lockdown risk reduction*” and “*lockdown reduction time*”. Similarly, the two variables respectively explain efficacy and starting time of lockdown policies in minimizing the flow of COVID-19. The SEIR model is sketched as seen in figure 1.

**Figure 1.**
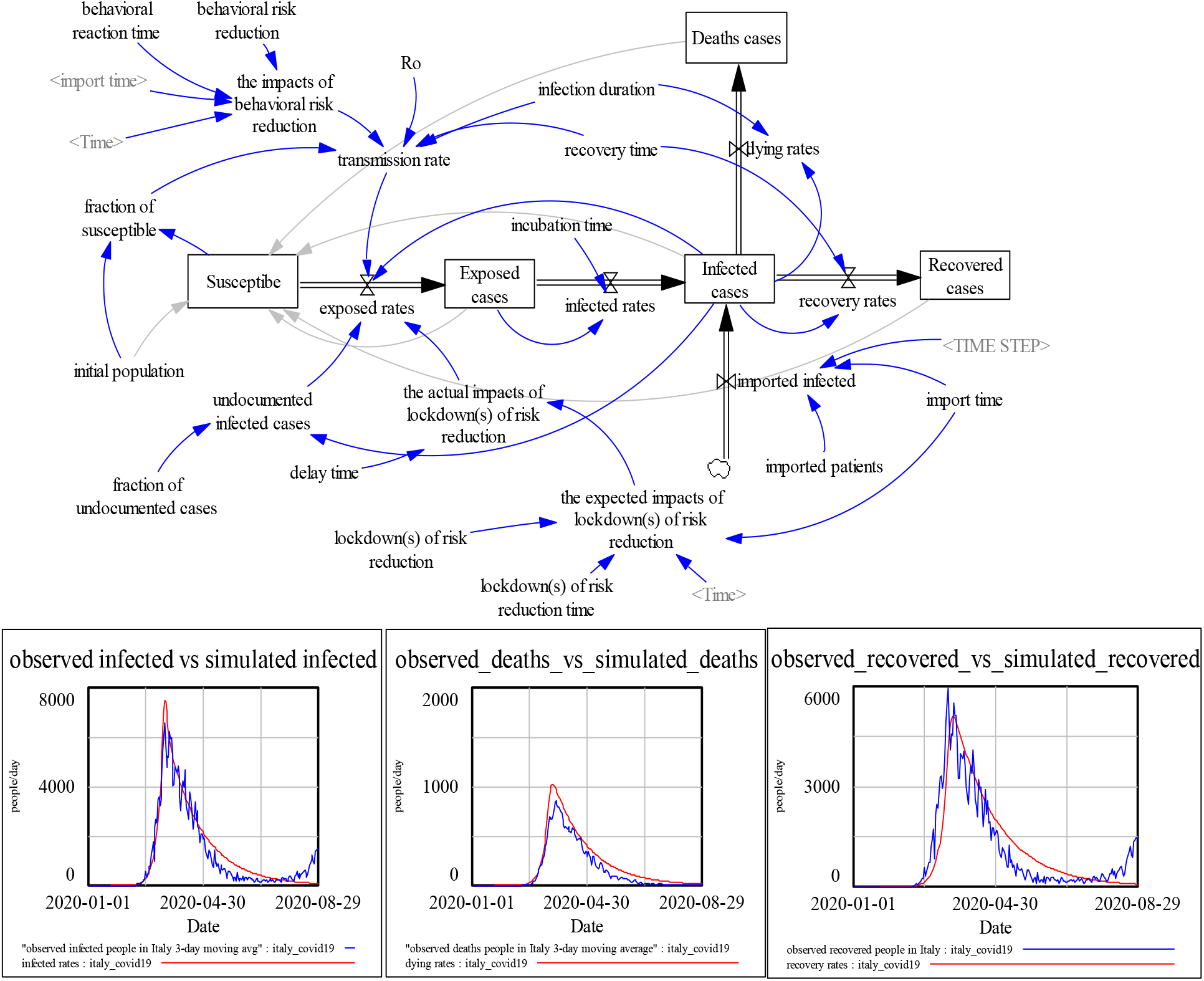
The SIER model for Italy

Other studies (Grosi et al., 2020; Goumenou et al., 2020) stated that undocumented cases may be responsible of rising cases in Italy and other cities in China (Li et al., 2020). The SDM reveals the roles of undocumented cases through two variables: “*undocumented infected cases*” and “*fraction of undocumented cases*”. A fraction of undocumented cases is a portion of undocumented cases compared with documented cases. In this model, documented cases are represented as “*infected rates*”.

As previously mentioned, the SEIR model separates patients into four categories, exposed, infected, dead, recovered, and undocumented cases. In the model, undocumented cases are represented as a multiplication between a fraction of undocumented cases and infected cases. Time variables such as recovery time is defined the average time between the symptom onset and recoveries while infected duration is defined as the average time between the symptom onset and deaths.

The SEIR model calculates the number of infected cases, deaths, and recoveries based on equations 1-3 as follows:

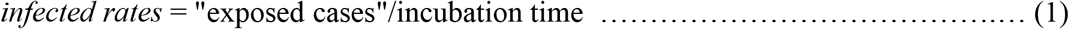

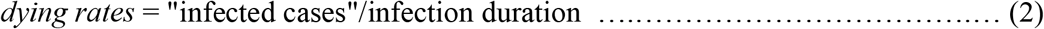

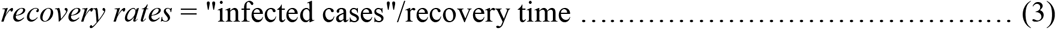

Transmission rate measures the number of exposed people after contacting or stand closes with infected people. Following Fiddaman (2020), transmission rate is calculated based on equation 3. For the first policy, its impact is calculated based on equation 4. Equation 4 means that the first policy of behavioral reduction risk decreases transmission rate based on two factors: “behavioral reduction risk” and “behavioral reduction time”.

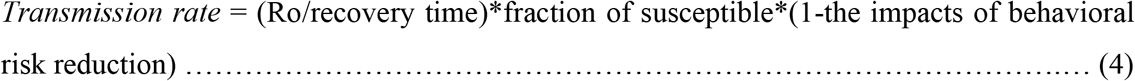

The second policy i.e. lockdowns is calculated similar to equation 4. Equations 5a and 5b show the number of exposed cases decreases after the second policy starts at “lockdown reduction time”.

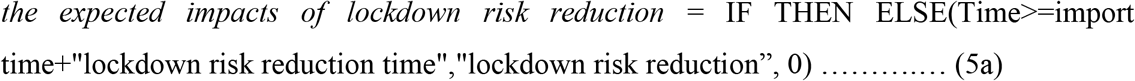

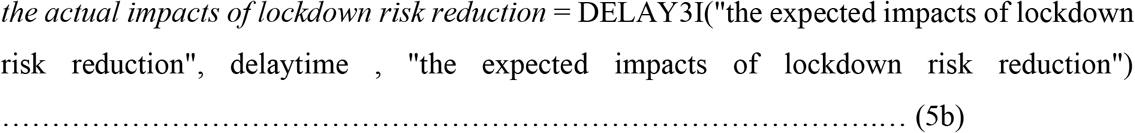

After running the calibration process, optimized values for each parameter are shown in table 2. The second and third columns represent optimized values if the first case is around January and around February 2020 respectively. As seen in table 2, there is no much differences between the two types of the first confirmed cases. This implies that the first policy i.e., voluntary acts such as social distancing and handwashing hindered the COVID-19 in early January 2020. In other words, the first policy was effective from January 31^st^, 2020 to February 21^st^, 2020.

**Table 2.** The best parameter values. Variables with asterisk should be quantified from the first confirmed case.

| No | Variables | Estimated values #1 | Estimated values #2 |
| --- | --- | --- | --- |
| 1 | Ro (basic reproduction number) | 3.68 (3-3.7) | 3.7 (3-3.7) |
| 2 | Incubation time (days) | 3 (3-5) days | 3 (3-5) days |
| 3 | Infection duration (days) | 16.5 (7-45) days | 12.56 (7-45) days |
| 4 | Recovery time (days) | 5.7 (3-35) days | 2.75 (3-35) days |
| 5 | Fraction of undocumented cases | 73% (46-62) % | 79% (46-62) % |
| 6 | The behavioral reaction time (days) | 37.87 days* | 20.7 days* |
| 7 | The behavioral risk reduction | 37.33% | 31.37% |
| 8 | The lockdown(s) of risk reduction time | 33 days* | 18 days* |
| 9 | The lockdown(s) of risk reduction | 65% | 65% |
| 10 | Delay time (days) | 5 days | 5.3 days |
| 11 | Import time | 31 days (the first confirmed cases should be a case announced on January 31<sup>st</sup>, 2020) | 52 days (the first confirmed case was February 21<sup>st</sup> or day 52 from January 1<sup>st</sup>, 2020) |

This study also shows that the first policy is helpful once the number of infected cases is relatively low (Piguillem & Shi, 2020). However, that study (Piguillem & Shi, 2020) also claimed that once the COVID-19 cases rise significantly so separating infected and disinfected people owing to is relatively difficult, policymakers must apply the second policy.

Looking back at table 2, it appears that voluntary acts such as handwashing and social distancing, despite their importance in hindering the COVID-19 flow, have the relatively lower effect than the lockdown policy (31%-37% vs 65%). However, this does not mean that voluntary acts are not needed. The first policy such as handwashing and social distancing could hinder the flow of COVID-19 in the first wave despite its relatively low effects.

This study also highlights that the important roles of undocumented cases, so-called asymptomatic cases, during the first wave of COVID-19 in Italy. This study finds that undocumented case was about 70% of the confirmed cases in Italy.

Moreover, the SEIR model can reproduce similar outputs compared to respective observed outputs as seen in figure 1. The SD model has symmetric Mean Percentage Errors (sMAPE) less than 10% as seen in table 3.

**Table 3.**
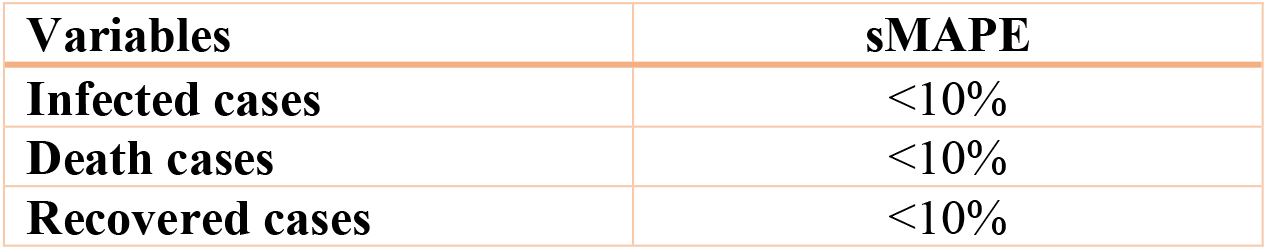
sMAPE for the SEIR model

| **Variables** | **sMAPE** |
| --- | --- |
| **Infected cases** | <10% |
| **Death cases** | <10% |
| **Recovered cases** | <10% |

## IV. Concluding remarks

Using the system dynamics approach, Italian COVID-19 is successfully modelled. Differing from other studies, this study aims to model dynamics of infected, recovered, and dead cases incorporating undocumented cases. Moreover, the SDM captures the positive effects of policy measures including behavioral policies (masks, handwashing, and social distancing) and lockdowns or isolations

Once optimized values for each parameter are obtained through the MCMC calibration process, the SEIR successfully reproduces similar outputs as observed outputs. The SEIR model also provides evidence that the first policy has relative lower impacts than the second policy. Nevertheless, the first policy is important once the number of infected cases are relatively low. Once the number of infected cases is high, the second policy shows relatively higher impacts in minimizing the flow of COVID-19. The higher impacts of the second policy than the first policy may be a clue to the second wave of the COVID-19 around the world.

*The SEIR model is available online at: https://osf.io/vuypf

## Data Availability

observed data are available in vdf file in the model

